# Impact of expanded influenza vaccination programs in children and adults in Australia: a mathematical modelling study

**DOI:** 10.64898/2026.11.23.26368629

**Authors:** Tiffany Leung, Isaac J. Stopard, James G. Wood, Alexandra B. Hogan

**Author notes:** Corresponding author: Dr Alexandra Hogan, UNSW Sydney, Sydney, NSW 2052, Australia.

## Abstract

**Background:** Seasonal influenza is a common cause of severe illness, particularly in young children and older adults. In Australia, influenza vaccination programs using the inactivated influenza vaccine (IIV) largely focus on direct protection of older adults. Live-attenuated influenza vaccine (LAIV) is now available for children aged 2–17 years, offering an opportunity to complement existing IIV programs.

**Methods:** We developed a mathematical model of seasonal influenza transmission and vaccination in Australia, calibrated to surveillance data. We evaluated the health impact of LAIV programs on influenza burden relative to a baseline IIV-only scenario that assumed current coverage levels. We compared the dose efficiency of LAIV programs targeting school-aged children with strategies that increased IIV uptake in adults.

**Findings:** An LAIV program targeting children aged 5–17 years was estimated to have the largest population-level impact, reducing influenza hospitalisations by 16% and deaths by 13%, assuming 55% coverage. An IIV program achieving 55% coverage in adults aged 50 years and older was predicted to have a similar population-level reduction in hospitalisations to a primary school-aged LAIV program (around 9.5%). Based only on doses administered, LAIV programs were more efficient at averting cases, hospitalisations, and deaths than strategies that expanded IIV uptake in adults. However, for outcomes in adults *≥*65 years old, increasing IIV uptake in this age group was estimated to be the most efficient strategy, with a number needed to vaccinate of 1811 to avert one hospitalisation.

**Interpretation:** Our results support the potential use of LAIV as an additional tool in the prevention of seasonal influenza in Australia.

**Funding:** NHMRC

**Research in context:** *Evidence before this study:* Annual influenza vaccination is recommended for everyone aged 6 months and older in Australia, but coverage remains suboptimal and is lowest among children aged 6–17 years. In 2026, an intranasally administered live-attenuated influenza vaccine (LAIV) became available for children aged 2–17 years, providing an alternative to the injectable inactivated influenza vaccine (IIV). School-based delivery of LAIV could increase vaccine uptake in children and reduce influenza disease burden across the population. We searched PubMed for studies published between January 1, 2000, and August 31, 2026, using the search terms “vaccin*” and (“mathematical model*” OR “transmission model*”) and (“estimat*” OR “evaluat*”) and (“influenza” OR “seasonal influenza” OR “live-attenuated influenza”), with no language restrictions. The search returned 277 records, of which 48 were population-level mathematical modelling studies that estimated the health impact of vaccination strategies against seasonal influenza. We identified two additional studies outside the search but known to the authors, bringing the total to 50 relevant studies. Of these, 31 were conducted in European settings, eight in North America, 10 in other or generic settings, and one specifically in Australia. The Australian study evaluated the health and economic benefits of increasing influenza vaccine coverage in children in Western Australia. Thirteen studies modelled LAIV as part of the vaccination strategy, predominantly in European settings. These studies evaluating LAIV programs generally indicated that vaccinating children could reduce influenza burden through both direct protection and indirect protection of non-target age groups, particularly of older adults. However, we identified no modelling study that compared the population-level impact of a school-aged LAIV program in Australia to strategies that increased influenza vaccine uptake in older adults.

*Added value of this study:* We developed a dynamic seasonal influenza transmission model calibrated to Australian surveillance data to compare the health impacts of LAIV programs targeting primary school-aged or all school-aged children with strategies that increased IIV uptake in adults aged 50–64 years or ≥65 years. At 55% coverage, we estimated that an LAIV program targeting children aged 5–17 years would result in the largest reductions in influenza cases, hospitalisations, and deaths due to influenza, both across the population and among adults aged ≥65 years. We found that a large proportion of outcomes averted through childhood vaccination occurred outside the target age group, demonstrating substantial indirect protection. In contrast, increasing IIV uptake in older adults, who are at higher risk of severe influenza, predominantly provided direct protection to that age group. We found that school-aged LAIV programs would likely require fewer vaccinations to avert a case, hospitalisation, or death across the population than strategies that increased IIV uptake in adults. However, for outcomes in adults aged ≥65 years, increasing IIV uptake in this age group required the fewest vaccinations per outcome averted.

*Implications of all the available evidence:* Our findings support the potential for school-aged LAIV programs, such as through school delivery, to complement existing IIV programs that focus on direct protection to older adults in the prevention of seasonal influenza in Australia. With the decline of influenza vaccination coverage since the COVID-19 pandemic, any modification to existing immunisation programs must consider programmatic feasibility, acceptability, and attainability for differing age groups.

## Introduction

Seasonal influenza is a common acute respiratory infection that causes 3 to 5 million annual cases of severe illness globally [1]. In Australia, while influenza burden varies by year, seasonal influenza can result in an estimated 18,000 hospitalisations and 3,500 influenza-associated deaths annually [2], with annual observed deaths recorded as due to influenza ranging from approximately 400 to 1300 in recent years [3, 4]. Children aged under 5 years and adults over 65 years are among those at higher risk of severe influenza-associated illness [2].

Despite recommendations for influenza vaccination for people over 6 months of age [5], coverage in Australia with the inactivated influenza vaccine (IIV) remains suboptimal at around 30% in recent years [6], with the lowest level of age-specific coverage in children aged 6–17 years, at around 12% [7]. In 2026 an intranasally administered live-attenuated influenza vaccine (LAIV) became available in Australia for children aged 2–17 years [8]. This needle-free vaccine option serves as an alternative to the traditional injectable IIV and may provide a means to increasing influenza vaccine coverage in children.

Children are known to play a key role in influenza transmission, particularly whilst at school [9, 10, 11], meaning that increasing vaccination in this age group is expected to yield both direct and broader population-level benefits. While newly available in Australia, LAIV has been used in Northern Hemisphere countries for over two decades. In the United Kingdom (UK), for example, school-based LAIV programs have been introduced in a phased approach since 2013. In the 2014–15 influenza season (the second season in which school-based LAIV programs were piloted), general practitioner consultations, hospital admissions, and respiratory swab positivity were consistently lower in pilot areas compared to non-pilot areas [12]. Vaccine uptake exceeded 50% in eligible children in 2015/16 [13, 14].

In this study we use a mathematical model to estimate the potential age-specific and population-level impact of a school-aged LAIV program in Australian children, relative to the existing IIV-only program, and assuming rollout timing and uptake that could potentially be achieved via school delivery. We compare the estimated health impacts of LAIV programs with that of expanded IIV programs that target adults under a range of different realistic age-targeting and coverage assumptions.

This work was conducted as part of a multi-model exercise conducted by the Australia–Aotearoa Scenario Modelling Hub.

## Methods

### Influenza transmission model

We developed an age-structured, deterministic compartmental model of seasonal influenza transmission and vaccination (Figure 1A). In the model, the population is divided into nine age groups: 0–*<*0.5, 0.5–*<*1, 1–4, 5–11, 12–17, 18–49, 50–64, 65–79, and 80+ years. For each age group, individuals are stratified by infection status: susceptible, exposed but not yet infectious, infectious, and recovered. Infection status is then further subdivided by vaccination status: unvaccinated without prior immunity, unvaccinated with prior immunity, and vaccinated.

**Figure 1:**
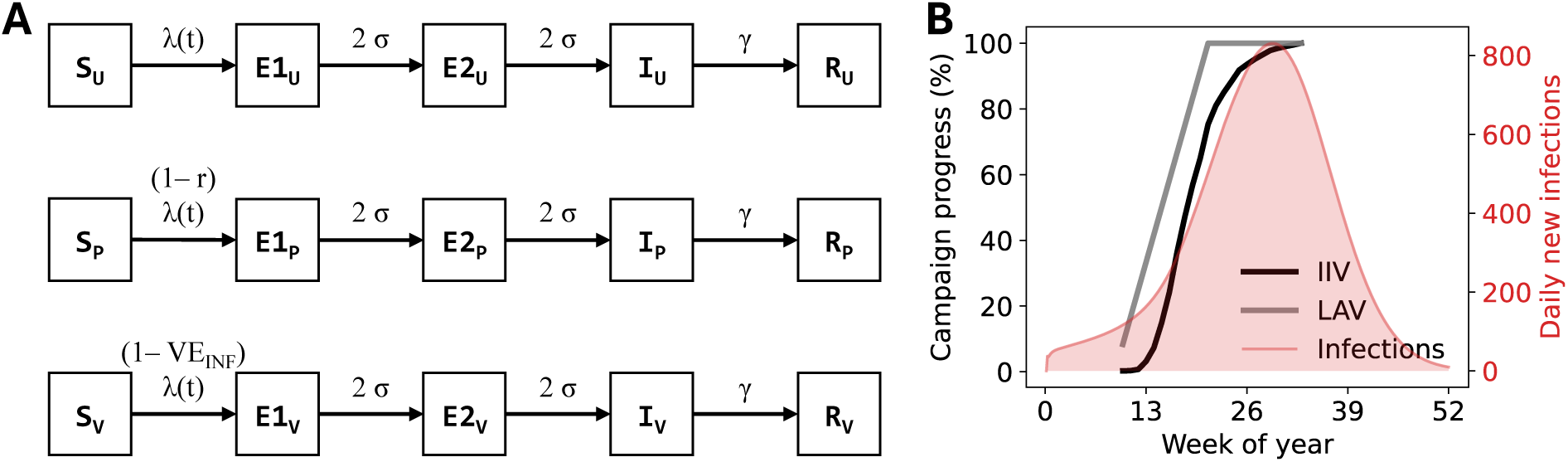
Model diagram of influenza transmission and vaccination campaign. (A) The population is divided according to disease status (susceptible, exposed but not yet infectious, infectious, and recovered, denoted by *S*, *E*, *I*, *R*) and vaccination status (unvaccinated without prior immunity *U*, unvaccinated with prior immunity *P*, and vaccinated *V*, denoted by subscripts), with transitions between compartments denoted by horizontal arrows. We assume that those who are vaccinated or who have prior immunity have a proportional reduction in susceptibility to infection, and that after infection, individuals recover and are fully immune. We assumed a gamma-distributed latent period by using a two-step transition from *E* to *I*. See Supplementary Material for model detail and equations. (B) The vaccination campaign progression for IIV (black line) and LAIV (gray line) are shown as a percentage relative to the total coverage for that scenario (left-hand y-axis) with one baseline infection incidence trajectory (pink line and shaded region; right-hand y-axis).

Seasonality in transmission as demonstrated by strong winter influenza epidemics is captured using a sinusoidal forcing function. We assumed no waning of infection-or vaccine-induced immunity over the modelled one-year time horizon, consistent with studies that found no evidence of waning of vaccine-induced protection over one season [15, 16]. We assumed that prior immunity conferred partial protection against infection and that this partial protection increased linearly over the first 18 years of life (from 0% at first to 21% at age 18 and uniform thereafter [17]).

### Setting

Yearly population sizes for the model calibration were obtained from the Australian Bureau of Statistics (ABS) [18]. We used an age distribution informed by Australian demographic data [18] and UK POLYMOD contact mixing patterns [19, 20].

To account for uncertainty in population-level prior immunity, we systematically varied the initial proportion of partially susceptible individuals due to prior immunity (0.05–0.75) and unvaccinated infectious individuals (10*^−^*^8^–10*^−^*^4^), generating 40 distinct initial populations. The remaining population was susceptible without prior immunity, and we assumed that no individuals were vaccinated with the season’s IIV or LAIV at the start of the simulation. For each of the 40 initial populations, free parameters were re-calibrated to influenza notification data as described below.

In each re-calibrated set of simulations, we explored the effects of vaccination scenarios in a fixed population of one million.

### Influenza surveillance data and model calibration

We used Australian surveillance data to calibrate our model and produce a typical annual seasonal influenza epidemic. Briefly, we obtained laboratory-confirmed influenza cases from 2015 to 2025 for all states and territories from the National Notifiable Diseases Surveillance System (NNDSS) and grouped these data to the country level [21]. We combined these two data types (excluding years 2020 and 2021) to produce an agespecific influenza case rate. FluTracking is a participatory surveillance system that monitors self-reported symptoms and testing behaviour for respiratory infections, including for influenza. We used data on testing behaviour from these FluTracking surveys (Figure S1) to estimate the temporal trends in the probability that a person with symptoms would seek a test.

We calibrated modelled cases to weekly age-specific case notification data in Australia [21]. To do this, we calculated cases as infections scaled by the probability that that an infection would be symptomatic (81% [22]) and the age-specific probability that a person would seek a test (estimated from FluTracking [23]). We additionally introduced two scaling factors to health-seeking behaviour in children *<*18 years old and adults *≥*65 years old to account for biases in the FluTracking sample. Noting that COVID test-seeking behaviour in the FluTracking data has been found to be higher than nationally representative cross-sectional survey [24], and to account for potential age-specific differences in test-seeking behaviour, we introduced two scaling factors to adjust health-seeking behaviour in children *<*18 years old and adults *≥*65 years old. We fitted the model using a maximum likelihood approach, assuming weekly age-specific influenza cases were Poisson-distributed. We allowed key model parameters to vary, namely, the basic reproduction number *R*_0_, amplitude and phase shift of seasonal forcing, and scaling factors to health-seeking behaviour. We repeated this fitting procedure for all 40 sets of initial populations, resulting in 40 fitted parameter sets each with a distinct initial level of population prior immunity.

To estimate the risk of influenza hospitalisations, we obtained principal diagnosis data for 2018–19, 2022– 23, and 2023–24 from the Australian Institute of Health and Welfare (AIHW) National Hospital Morbidity Database (Figure S2A) [25]. Age-specific influenza deaths were estimated using data on deaths due to acute respiratory infections in Australia in 2023, 2024, and 2025 from the ABS (Figure S2B) [4]. We computed the number of hospitalisations and deaths from each infection trajectory using these age-specific risks of influenza hospitalisation and death respectively.

A full description of the data, model calibration, and methods is provided in the Supplementary Material.

### Vaccination strategies

We considered vaccination strategies that increased IIV coverage or introduced LAIV programs in different target age groups, compared to an IIV-only baseline in which coverage was assumed to be 55% for adults *≥*65 years old, 20% for adults 50–*<*65 years old, and 10% for children 5–*<*18 years old (Table 1):

- **IIV 65:** IIV coverage increased to 80% in adults *≥*65 years old;
- **IIV 50:** IIV coverage increased to 55% in adults 50–*<*65 years old;
- **LAIV 5–12:** LAIV introduced to children 5–*<*12 years old with 55% coverage;
- **LAIV 5–18:** LAIV introduced to children 5–*<*18 years old with 55% coverage.

**Table 1:** Vaccination coverage by age group and scenario. The vaccine product varies depending on the age group receiving the vaccine (adults aged 18 years and older always receive the IIV). Population does not sum to 100% due to rounding. Bolded values indicate where coverage differs from the baseline scenario.

|  |  | **Vaccination scenario** |  |  |  |  |
|---|---|---|---|---|---|---|
| **Age (years)** | **Population** | **Baseline** | **IIV 65** | **IIV 50** | **LAIV 5–12** | **LAIV 5–18** |
| 0–<0.5 | 1% | 0% | 0% | 0% | 0% | 0% |
| 0.5–<1 | 1% | 15% | 15% | 15% | 15% | 15% |
| 1–4 | 4% | 15% | 15% | 15% | 15% | 15% |
| 5–11 | 8% | 10% | 10% | 10% | **55%** | **55%** |
| 12–17 | 7% | 10% | 10% | 10% | 10% | **55%** |
| 18–49 | 44% | 20% | 20% | 20% | 20% | 20% |
| 50–64 | 17% | 20% | 20% | **55%** | 20% | 20% |
| 65–79 | 13% | 55% | **80%** | 55% | 55% | 55% |
| 80+ | 4% | 55% | **80%** | 55% | 55% | 55% |

For each scenario, non-target age groups received the standard IIV coverage rates at baseline. As a sensitivity analysis, we modelled IIV 50, LAIV 5–12, and LAIV 5–18 programs with coverage of the target ages reaching 30% or 80%. Note that in the IIV 50 high-coverage (80%) setting, adults *≥*50 years old would reach 80%, equivalent to IIV 65 plus IIV 50, while in the IIV 50 low-coverage setting, adults aged 50–64 years would reach 30% and adults *≥*65 years old would maintain the baseline coverage of 55%.

The baseline IIV coverage rates for different age groups were informed by the national influenza vaccination coverage data in Australia [7] (Table 1). For the IIV, we assumed 55% effectiveness against symptomatic disease in Australia [6]. We calculated 44% effectiveness against acquiring infection by assuming that the effectiveness against symptomatic disease in an infected person is 20% [26]. We set effectiveness against hospitalisation and death equal to that against symptomatic disease, as in Kelly et al. [27].

For the LAIV programs, we set an LAIV coverage of 55% in the target age group, consistent with data from the rollout of school-based programs in the UK [13, 14], and assumed a 65% vaccine effectiveness against symptomatic disease [28]. Using the same methods as for the IIV, we calculated a 56% effectiveness against acquiring infection and set a 65% effectiveness against hospitalisation for the LAIV. We used a ‘leaky’ vaccine that provided partial protection to all vaccinated individuals [29].

Rollout of both vaccines began in Week 10 of the year (start of March) (Figure 1B). The IIV was administered over 24 weeks, until Week 34 (end of August), based on data from the National Centre for Immunisation Research and Surveillance [7]. Assuming that the LAIV program is school-based and would be delivered before the peak influenza season, we simulated a 12-week rollout that lasted until Week 22 (end of May), within Terms 1 and 2 of the Australian school calendar [30, 31] (see Supplementary Material C).

### Model outcomes

For each scenario (and set of initial conditions and parameter values), we calculated the age-specific cumulative infections, cases of disease, hospitalisations, and deaths due to influenza, over one year from January to December. We calculated the number needed to vaccinate (NNV), a metric that represents the number of individuals that must be vaccinated in order to prevent one outcome (case, hospitalisation, and death due to influenza); it was calculated as the ratio of 1) the total additional doses required by each strategy compared to baseline and 2) the number of averted outcomes in all ages and in adults *≥*65 years old. We report the mean and range over 40 parameter sets.

## Results

### Model calibration

The fitted model broadly captured the seasonal and yearly variation in notified influenza cases: cases peaked during the middle of the year and increased annually from 2015 to 2025 (Figure S3). Baseline case trajectories were consistent with the confirmed influenza case data; calibration is illustrated in Figure 2A using case data for 2024, aggregated across ages groups and jurisdictions. We estimated a mean *R*_0_ of 1.28 (range 1.13–1.45) across the 40 sets of initial parameters, corresponding to a population-level mean infection attack rate of 12.2% (range 11.7–13.5%) at baseline. We found a strong correlation between the fitted parameters *R*_0_, the initial proportion with prior immunity, and the initial infection prevalence, indicating a lack of parameter value identifiability (Figure S5). Modelled cumulative hospitalisations and deaths at baseline aligned with observed influenza-attributable hospitalisations and influenza deaths (Figure 2B and C). For the full model calibration of influenza cases by year and age group, see Figures S3 and S4.

**Figure 2:**
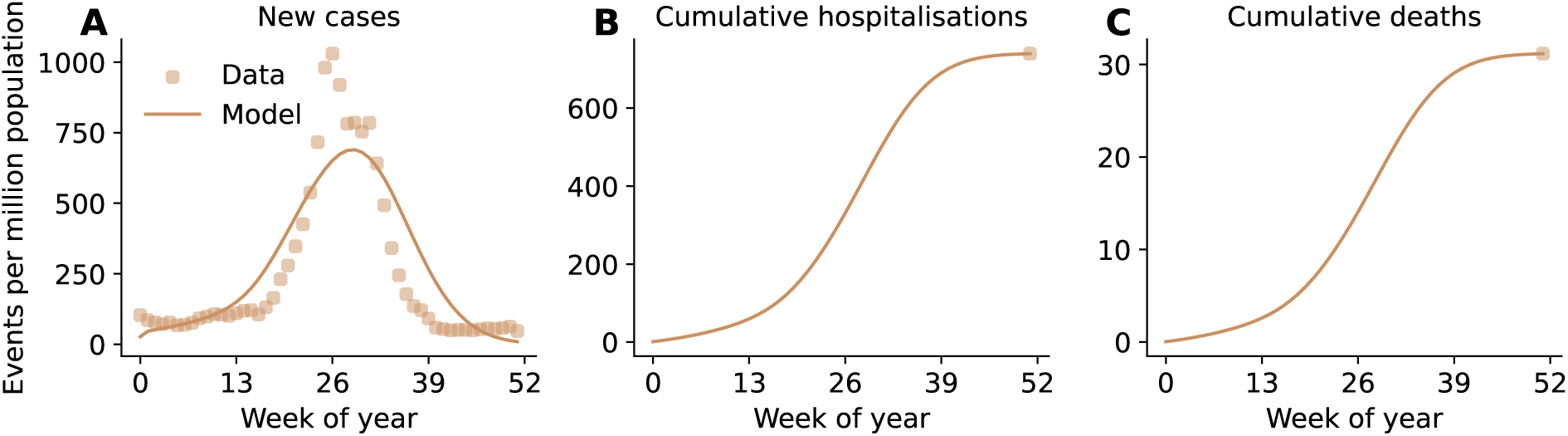
Modelled new cases of disease, cumulative hospitalisations, and cumulative deaths with calibration data over a one-year time horizon. One modelled baseline trajectory is shown alongside (A) weekly case data for 2024 from the NNDSS, aggregated across jurisdictions and age groups and scaled from the Australian population (27,387,790) to 1 million population; (B) estimated average annual influenza hospitalisations based on data from the AIHW; and (C) estimated average annual influenza deaths based on data from the ABS. Modelled cases are shown as weekly counts, whereas modelled hospitalisations and deaths are cumulative. Note that while the calibration to a single calendar year is illustrated here (2024, the most recent complete year of data), the model was fitted to age-specific data for nine years simultaneously. Figure S3 shows the model fitted to the age-specific case data for 2015–2025 (excluding 2020 and 2021), and Figure S4 shows the model fit with all ages aggregated. More details on the surveillance data are provided in Figure S2 and the Supplementary Material.

### Modelled baseline scenario

At baseline, we modelled a mean of approximately 122,000 (range 116,581–134,563) total infections per million population annually, resulting in approximately 15,000 (range 14,220–15,694) cases, 740 (range 737–742) hospitalisations, and 31.2 deaths (range 31.0–31.4) annually. These estimates correspond to approximately 14% of infections becoming cases, a 5% case-hospitalisation ratio, and a 0.2% case-fatality ratio (Table S2).

### Estimated impact on influenza hospitalisations and deaths

Of the modelled scenarios, an LAIV program targeting children aged 5–*<*18 years (LAIV 5–18) was estimated to have the largest reductions in hospitalisations and deaths at the population level, averting 16% (range 14–17%) of hospitalisations and 13% (range 10–14%) of deaths (Figure 3B, C). In contrast, an IIV program targeting adults aged 50–64 years with 55% coverage (IIV 50) and an LAIV program targeting primary schoolaged children (LAIV 5–12) each reduced hospitalisations by approximately 10%. Increasing coverage in adults *≥*65 years old (IIV 65) was estimated to achieve the smallest population-level reductions in hospitalisations at 4% (range 3.8–4.9%). For deaths at a population level, programs that target adults were predicted to lead to higher reductions than the LAIV 5–12 program; the IIV 50 program reduced deaths by 9.8% (range 7.1–12.8%), compared to 6.8% (range 5.2–7.6%) for LAIV 5–12 (Figure 3C).

**Figure 3:**
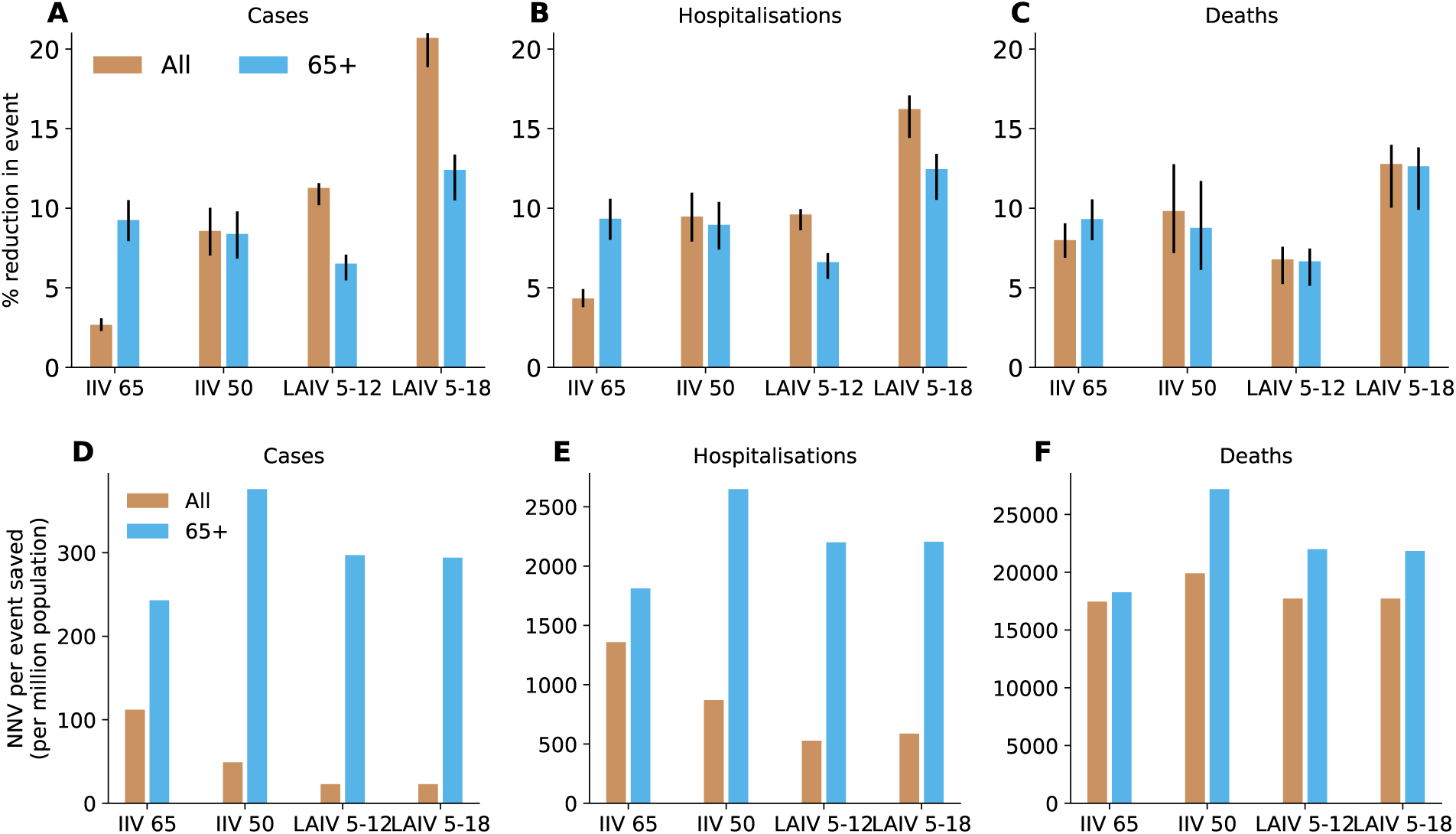
Mean percentage reduction and number needed to vaccinate (NNV) to prevent one outcome by strategy. Reductions in (A) cases, (B) hospitalisations, and (C) deaths relative to baseline; NNV to prevent (D) one case, (E) one hospitalisation, and (F) one death relative to baseline. The bars and black lines show the mean and range over 40 simulations. Figure S6 shows the number of IIV and LAIV doses used per strategy.

When the outcomes were restricted to events averted in adults *≥*65 years old, we found that programs targeting adults had a greater impact than LAIV 5–12: IIV 65 and IIV 50 each reduced hospitalisations by approximately 9%, compared to 6.6% for LAIV 5–12 (Figure 3B). An LAIV program targeting children aged 5–*<*18 years was still predicted to have the largest impact, reducing hospitalisations and deaths by 12.5% (range 10.5–13.4%) and 12.6% (range 9.9–13.8%) respectively (Figure 3B and C).

The health benefits in target and non-target age groups differed for each vaccination program, reflecting their direct and indirect impacts. Programs targeting older adults provided more direct protection, whereas those targeting children provided more indirect protection. For example, when coverage in adults *≥*65 years old increased to 80% (IIV 65), 96% of averted deaths were in the target age group (Table S3). In contrast, only 3% of the deaths averted were in the target age group in an LAIV program targeting children aged 5–*<*18 years, showing that most deaths were averted through indirect protection. A similar pattern was observed for averted hospitalisations, with larger direct impact when targeting adults *≥*65 years old and greater indirect impact from targeting children (Table S3).

### Estimated impact on influenza cases

At the population level, we estimated that LAIV programs would have the largest impact on cases, with LAIV 5–18 averting cases by 20.7% (range 18.9–21.7%), while LAIV 5–12 averted 11.3% (range 10.2–11.6%) (Figure 3A). In comparison, we estimated that increasing IIV coverage from 55% to 80% in adults *≥*65 years old (IIV 65) would have the smallest population-level impact on cases, reducing by 2.7% (range 2.3–3.1%).

### Efficiency of the modelled influenza vaccine strategies

We calculated the number needed to vaccinate (NNV) to measure the efficiency of the modelled scenarios. Across all ages, the school-aged LAIV programs had lower NNVs than expanded IIV programs that targeted adults for preventing cases or hospitalisations, whereas a program targeting adults *≥*65 years old (IIV 65) had the lowest NNV for preventing deaths (Table 2). For example, the NNV to prevent one hospitalization in all ages was 527 for an LAIV program targeting primary school-aged children (LAIV 5–12), compared to 1358 for IIV 65 (Table 2). The NNVs were similar between the two LAIV programs across all outcomes, with LAIV 5–12 slightly more efficient than LAIV 5–18. For example, 527 vaccine doses were required to prevent one hospitalisation for LAIV 5–12, compared to 588 doses for LAIV 5–18, a difference of 61 doses (per million population).

**Table 2:** Averted outcomes, total vaccine doses, and number needed to vaccinate to avert 1 case, 1 hospitalisation, and 1 death in all ages and in adults aged 65 and older. (compared to baseline of 241,639 doses per million population). The mean over 40 simulations is presented.

| **Scenario** | **Averted**<br>**cases** | **Averted**<br>**hospitalisations** | **Averted**<br>**deaths** | **Total**<br>**extra doses** | **NNV to avert**<br>**one case** | **NNV to avert**<br>**one hospitalisation** | **NNV to avert**<br>**one death** |
|---|---|---|---|---|---|---|---|
| **In all ages** |  |  |  |  |  |  |  |
| IIV 65 | 388 | 32 | 2.49 | 43,469 | 112 | 1,358 | 17,457 |
| IIV 50 | 1,245 | 70 | 3.06 | 60,914 | 49 | 870 | 19,907 |
| LAIV 5–12 | 1,637 | 71 | 2.11 | 37,388 | 23 | 527 | 17,719 |
| LAIV 5–18 | 3,008 | 120 | 3.98 | 70,539 | 23 | 588 | 17,723 |
| **In 65+** |  |  |  |  |  |  |  |
| IIV 65 | 179 | 24 | 2.38 | 43,469 | 243 | 1,811 | 18,264 |
| IIV 50 | 162 | 23 | 2.24 | 60,914 | 376 | 2,648 | 27,194 |
| LAIV 5–12 | 126 | 17 | 1.70 | 37,388 | 297 | 2,199 | 21,993 |
| LAIV 5–18 | 240 | 32 | 3.23 | 70,539 | 294 | 2,204 | 21,839 |

When the NNV was restricted to adults *≥*65 years old, increasing coverage in this age group (IIV 65) directly was the most efficient strategy, with NNVs of 1811 to prevent one hospitalisation and 18264 to prevent one death (Table 2). Both school-aged LAIV programs had lower NNVs (and hence were more efficient) than a program that increased coverage in adults aged 50–64 years.

### Sensitivity analysis

We explored the impact of achieving a lower (30%) and higher (80%) coverage in the target age groups for LAIV 5–12, LAIV 5–18, and IIV 50, compared to the assumed default coverage of 55%. At 30% coverage, the LAIV 5–18 program continued to have the largest population-level reductions for all outcomes, reducing hospitalisations and deaths by 9% and 7% respectively, compared to baseline (Table S3).

At 80% coverage, with the IIV 50 target age expanded to adults *≥*50 years old (from adults aged 50–64), we estimated LAIV 5–18 to have the highest reductions in infections (23%, range 20.8–24.4%), cases (29%, range 27–30.8%), and hospitalisations (23%, range 20.6–24.2%) (Table S3). In contrast, IIV 50 had the largest reduction in deaths, at 22% (range 19.3–24.9%) compared to baseline.

The efficiency of the vaccination programs differed slightly at lower (30%) and higher (80%) coverage in the target age groups. At both coverage levels, the school-aged LAIV programs were more efficient at preventing all three outcomes, both across all ages and among adults *≥*65 years old (Table S4). This differed from the default 55% coverage scenarios, in which directly increasing IIV coverage in adults *≥*65 years old (IIV 65) was the most efficient strategy for preventing outcomes in that age group.

## Discussion

In this modelling study we estimated that an LAIV program targeting primary and secondary school students and achieving 55% coverage would have larger population-level benefits, reducing more influenza cases, hospitalisations, and deaths, compared to strategies that expanded IIV uptake in adults. School-aged LAIV programs were also estimated to achieve the greatest reductions in these outcomes among adults *≥*65 years old, highlighting the substantial indirect protection provided by vaccinating school-aged children. When accounting for the number of vaccine doses administered, we estimated that at the population level, LAIV programs would generally be more efficient than strategies that increased IIV uptake in older adults. However, if we focused on measuring prevented outcomes in adults *≥*65 years old, increasing IIV coverage to 80% in this age group was predicted to be the most efficient strategy.

While modelled health impact and efficiency are important considerations for influenza vaccination program policy, programmatic feasibility is also relevant. Australia has an established infrastructure that delivers vaccines through general practices, pharmacies, and residential aged care facilities, as well as school-based programs for diseases such as human papillomavirus [32]. However, uptake of influenza vaccination has declined across all age groups since the COVID-19 pandemic [33]. Hence, achieving 55% coverage (as assumed in our scenarios) would represent a substantial increase from the current uptake in Australian school-aged children. However, coverage exceeding 50% has been achieved through school-based programs that delivered intranasal LAIV in the UK and other European countries [13, 14], suggesting that this level of uptake may be feasible with school-based delivery. The barriers to increasing uptake are multiple, and include practical considerations (ease of access and cost), as well as behavioural and social factors such as perceived autonomy in vaccine decision-making, trust in health workers, and community leader influence [34]. Any modifications to existing influenza vaccine programs would need to consider acceptability and achievable uptake for differing population groups.

Compared to the IIV-only baseline scenario (with 24% coverage across the population), we found that an LAIV program achieving 55% coverage in children aged 5–17 years would reduce influenza cases by 21%, hospitalisations by 16%, and deaths by 13% across the population. Our findings are broadly consistent with those of de Boer et al. [35], who modelled LAIV vaccination in children aged 2–16 years in the Netherlands. Assuming 50% coverage, they estimated an average annual population-level reduction of 18% in cases, 7% in hospitalisations, and 10% in deaths over 20 years, relative to a baseline vaccination program with approximately 21% coverage across the population. However, our modelled impact was smaller than in Wenzel et al. [36], who estimated that vaccinating 5–16-year-olds at 55% coverage would reduce cases, hospitalisations, and deaths by a mean of approximately 50% annually relative to their baseline scenario in England and Wales over 19 influenza seasons. One factor in their higher estimated reductions may be differences in vaccine efficacy against infection, which ranged from 28% to 70% in their model and varied by the degree of matching between vaccine and circulating strain and by age and risk group.

We estimated that an LAIV program achieving 55% coverage in children aged 5–*<*18 years would avert around twice as many cases as an IIV program achieving the same coverage in adults aged 50–64 years. Baguelin et al. [37] found a similar difference relative to their baseline scenario without vaccination: targeting adults aged 50–64 years reduced cases by around 28%, approximately half the 55% reduction achieved by targeting children aged 5–16 years (where impact was reported as the average events for each outcome across a range of coverage scenarios from 15–70%). Similar patterns were observed for severe outcomes. In our study, targeting adults aged 50–64 years averted 44% fewer hospitalisations and 23% fewer deaths than targeting children aged 5–*<*18 years. Baguelin et al. reported comparable relative differences, where vaccinating adults aged 50–64 years averted 39% fewer hospitalisations and 26% fewer deaths than vaccinating children aged 5–16 years [37].

We estimated that an LAIV program targeting children aged 5–17 years would provide substantial indirect protection, with around 97% of averted deaths and 66% of averted hospitalisations occurring in the nontarget age groups. Other modelling studies reported similarly high indirect benefits from vaccinating children against influenza [38, 39, 35, 40, 41]. For example, de Boer et al. [35] estimated that, for a program targeting children aged 2–16 years, on average, around 99% of averted deaths and 52% of averted hospitalisations occurred within the non-target age group, indicating a large proportion of severe outcomes were prevented due to indirect protection from vaccinating children. In contrast, we found that programs targeting older adults primarily provided direct protection to the target age group, with relatively little benefit outside the target age group. Together, these findings highlight the potential for LAIV programs that target school-aged children, which could be implemented through school-based delivery, to reduce severe disease in older adults (those at higher risk of severe influenza) through indirect protection.

In our study we estimated that an LAIV program in children would be more efficient (measured as vaccine doses administered) in terms of averted cases, hospitalisations and deaths compared to programs that increase coverage in adults aged 50–64 years. This finding is consistent with a study that modelled influenza transmission over 14 seasons in England and Wales [39]. Baguelin et al. compared strategies that either increased coverage to 50% in children aged 5–16 years or in adults aged 50–64 years and estimated greater reductions in cases and deaths, and greater dose efficiency, when increased coverage was targeted to children.

Our study has several limitations. First, we assumed that LAIV was delivered over a 12-week period before the epidemic peak. While a slower modelled rollout may have reduced the estimated impact, our assumption may be conservative, as school-based influenza vaccination programs in the UK occurred over 6–8 weeks [42]. We also assumed that the relative speed of IIV uptake was the same for all age groups. In reality, adults *≥*65 years old may be vaccinated earlier than other age groups [7]. Second, we assumed uniform vaccine efficacy across all age groups, and did not explicitly account for the use of different vaccines in adults *≥*65 years old. However, given the use of adjuvanted and high-dose influenza vaccines to address lower efficacy from standard vaccines in that age group [43], our efficacy assumption is likely to be reasonable.

Third, there was a paucity of high-quality population-based surveillance data to inform our assumptions around underlying influenza burden, particularly for non-symptomatic and symptomatic infections. Using a combination of data sources we inferred a mean annual attack rate of 12.2%, but noted considerable heterogeneity in age-specific influenza attack rates in the literature [44]. In addition, while we use Australian data to estimate severe influenza morbidity and death burden, these data are still subject to both ascertainment and under-reporting biases. Consequently, the absolute number of events averted by each vaccination strategy should be interpreted in the context of this uncertainty. Fourth, our model used age-specific contact patterns from POLYMOD [19]. Social mixing patterns may have changed since this study was performed, particularly following the COVID-19 pandemic. The recent Reconnect survey, conducted in the UK between November 2024 and March 2025, reported 20% fewer contacts on average than POLYMOD, with higher contact rates in children under 5 years old but lower contact rates in those above 5 years old [45]. Changes in these mixing patterns could influence transmission and the magnitude of indirect protection provided by childhood vaccination.

With the recent introduction of LAIV products in Australia, there is a need for evidence to inform decisions regarding program design, target age groups, and implementation. However, few mathematical modelling studies have quantified the impact of LAIV programs, and most are focused on European settings [39, 35, 36]. In this study, we compared the health impacts and efficiency of school-aged LAIV programs with strategies that increased IIV coverage in adults in Australia. Our findings provide timely evidence on the potential direct and indirect benefits of school-aged LAIV programs and the relative impacts of other adult-targeted IIV strategies, to support the design and evaluation of future influenza vaccination programs in Australia.

## Data Availability

All data produced in the present study are available upon reasonable request to the authors.

## Contributors

All authors meet the ICMJE criteria for authorship. ABH, TL, and JGW conceived and designed the study. TL and IJS developed the model, with input from ABH and JGW. IJS conducted the model calibration, and TL performed the scenario simulations and generated the results. ABH and JGW were responsible for funding acquisition. All authors contributed to the interpretation of results. TL wrote the manuscript, and all authors reviewed and edited it. All authors approved the final version of the submitted manuscript, had access to the data used in this study, and had final responsibility for the decision to submit for publication.

## Acknowledgements

The authors thank Freya M Shearer, Oliver Eales, Eliran Boksenbojm and James M McCaw for providing helpful insights on this study as part of the Australia–Aotearoa Scenario Modelling Hub multi-model exercise. We thank the ACEFA peer reviewers, Kathryn Glass, Nick Golding, and Michael J Plank. We thank Alys R Young, Katharine L Senior and Ruarai Tobin for contributions to data processing and curation, and Kristine Macartney, Allen C Cheng and Bette Liu for valuable insights on interpretation of data and model outputs. We acknowledge individuals who contributed to the collection and curation of datasets used in this study, that were provided by the National Notifiable Diseases Surveillance System, FluCAN, and FluTracking, and thank community participants of the FluTracking system. National Notifiable Disease Surveillance System data were provided by the Interim Australian Centre for Disease Control, Australian Government Department of Health and Aged Care, on behalf of the Communicable Diseases Network Australia (CDNA). We thank public health staff and data managers in state and territory health departments, and the Australian Government Department of Health and Aged Care, along with state and territory public health laboratories.

## Declaration of generative AI and AI-assisted technologies

The authors used ChatGPT (GPT-5.5) for language editing and improving clarity and readability. Google Gemini was used to help with minor debugging of code. The authors reviewed and edited the output as needed and take full responsibility for the content of the manuscript.

## Supplemental Material

## A Supplementary Figures

**Figure S1:**
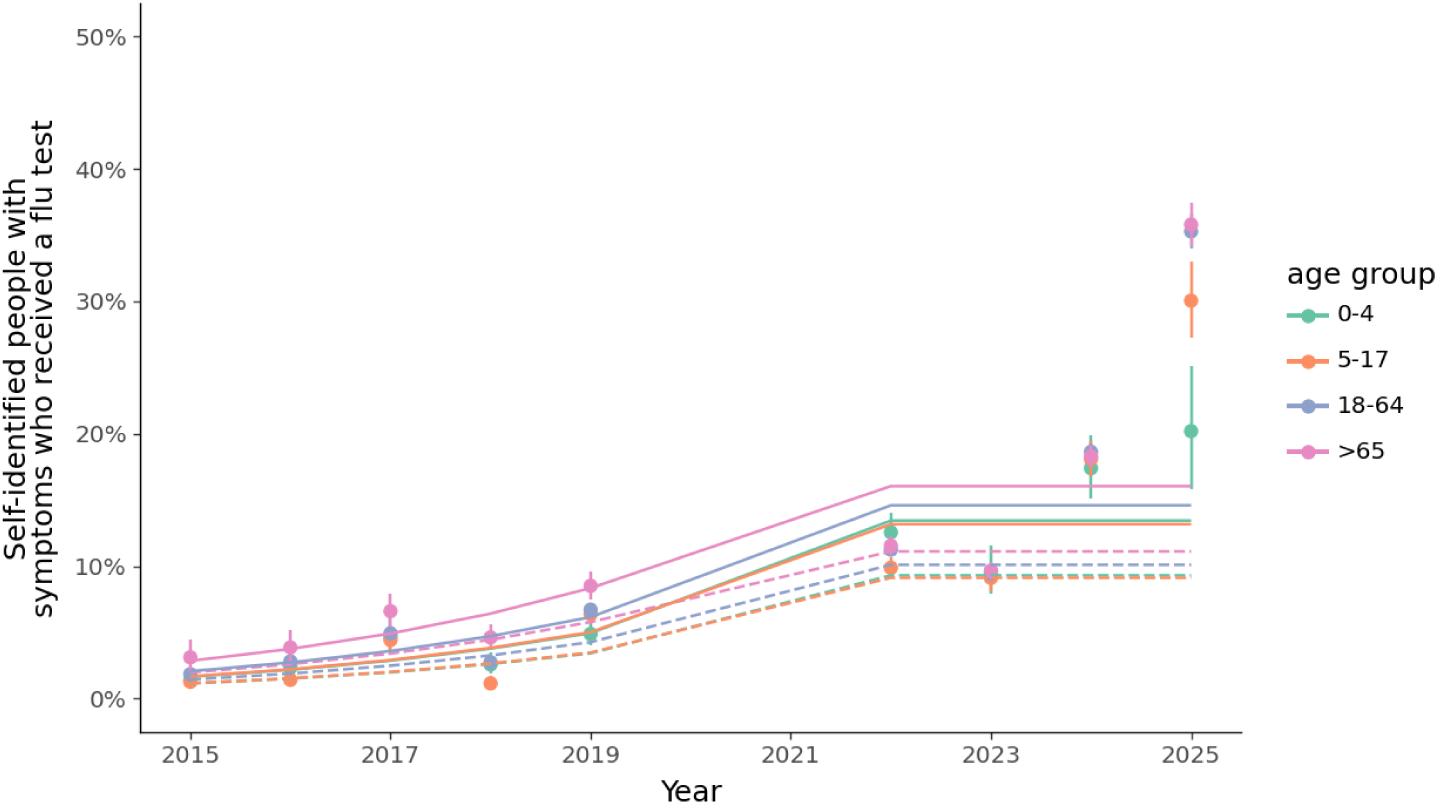
Age- and year-specific proportion of reports of individuals with at least one influenza symptom who also received an influenza test, calculated using data sourced from the FluTracking surveillance system, for each calendar year. The colour indicates the age group. The points indicate the sampled values and the solid lines indicate the modelled values. The dashed lines show the adjusted values taking into account differences between the FluTracking data and national behavioural survey, which were used in the transmission model calibration. Data for the 2020 and 2021 calendar years were excluded due to the COVID-19 pandemic. Note that we made no adjustment for the type of test (rapid antigen or polymerase chain reaction) and that the composition of test types changed over time.

**Figure S2:**
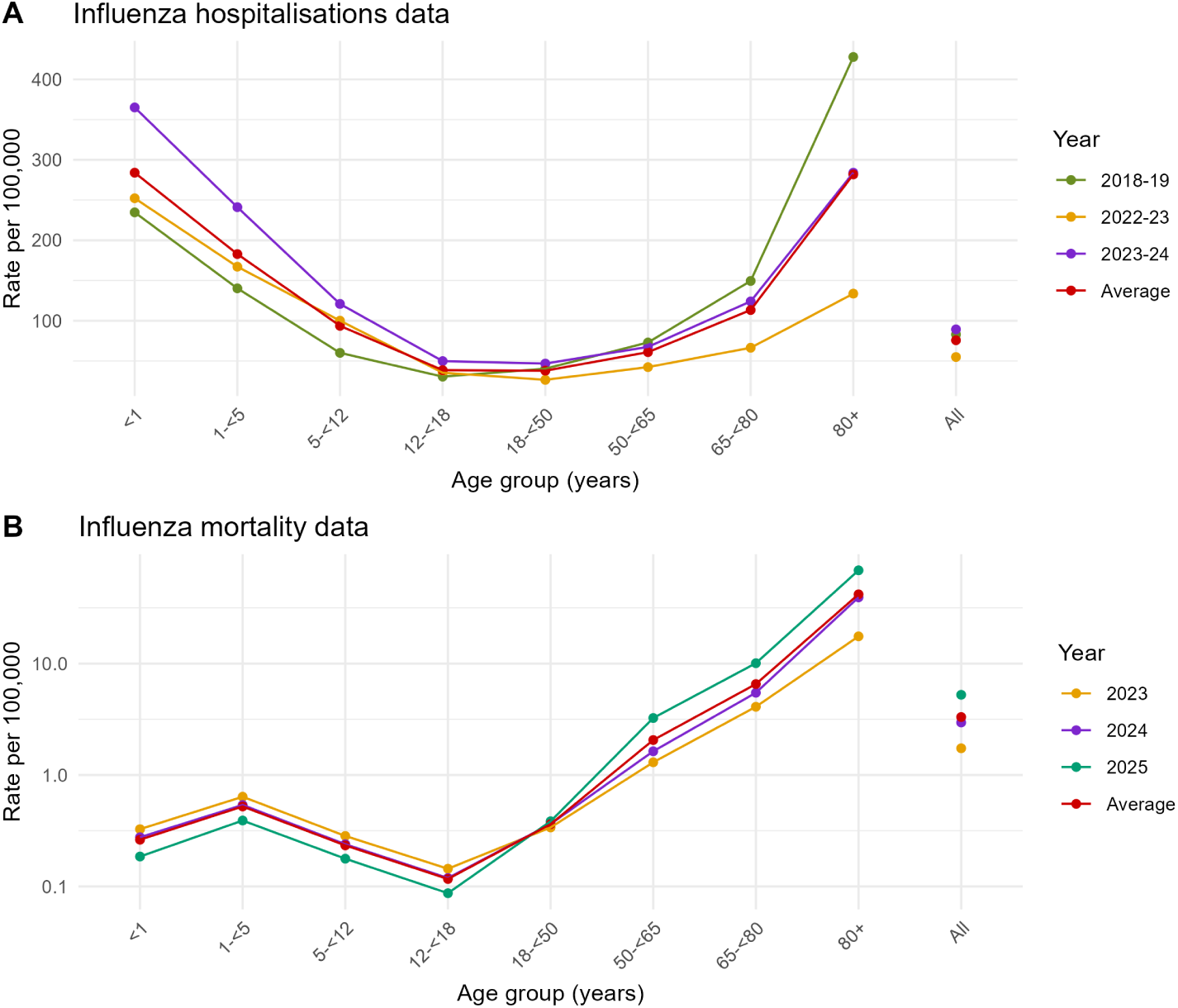
Age- and year-specific influenza hospitalisation and mortality data. (A) Influenza hospitalisation data were extracted from the Australian Institute of Health and Welfare (AIHW) Principal Diagnosis data cubes (codes J09, J10, J10.1 and J10.8) for three financial years (2018–19, 2022–23 and 2023–24) [25]. We used the corresponding Australian Bureau of Statistics population size for each year to estimate the age-specific hospitalisation rate for each year, and calculated the average rate (shown in red points and line) to use in our analysis. (B) Age-specific influenza deaths for age groups 0–19 years, and in 10-year age groups thereafter, for years 2023, 2024 and 2025 were obtained from the Australian Bureau of Statistics [4] and converted to age-specific rates using the corresponding population size for each year. To estimate the age-specific rates in children younger than 19 years, we applied the age-specific proportions of influenza deaths in child age groups for 2018–23 reported in McRae et al. 2025 [46]. The average rate across the three years (shown in red points and line) was used in our analysis.

**Figure S3:**
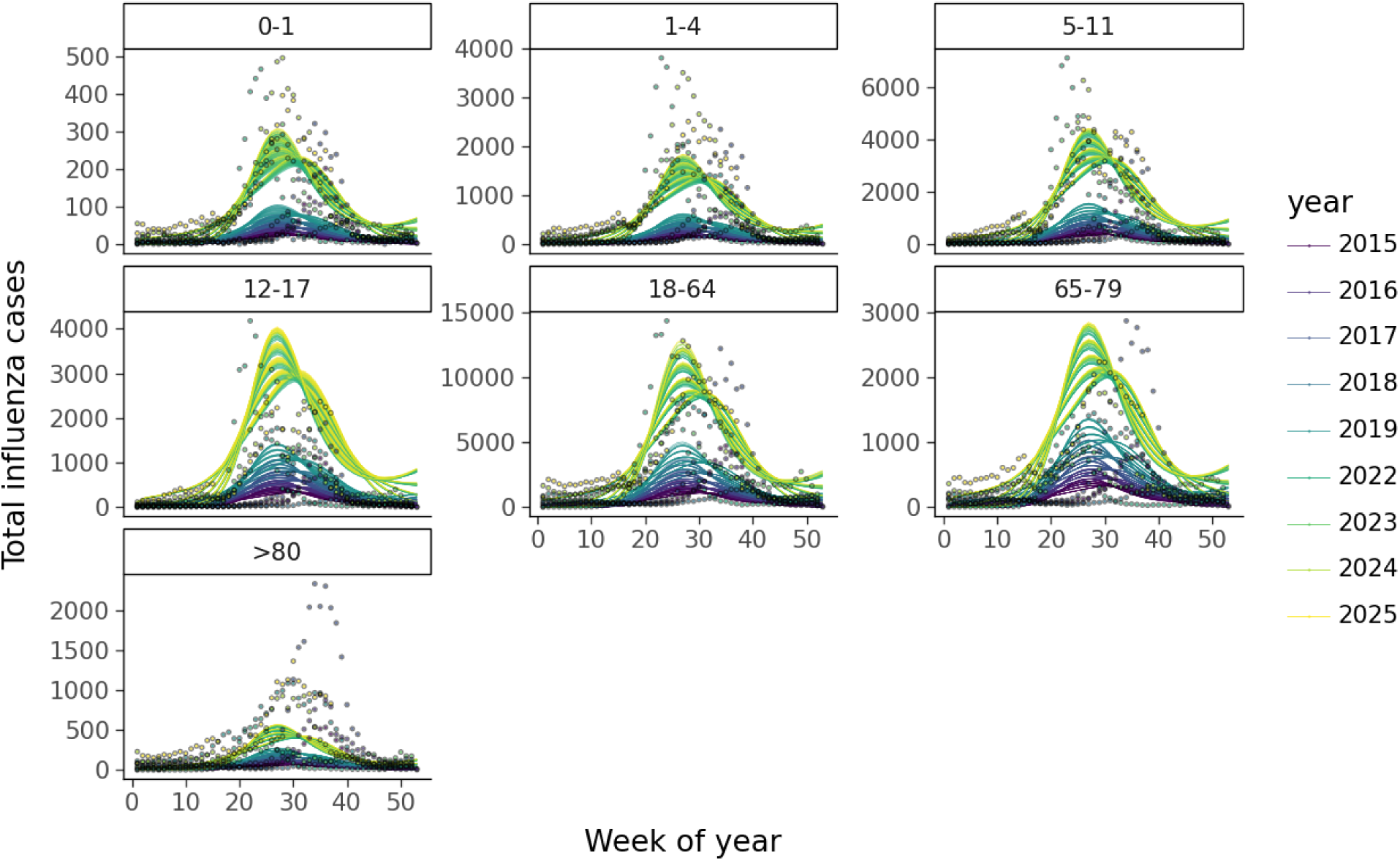
Model calibration. The number of cases was obtained from the Australian National Notifiable Diseases Surveillance System. The colour indicates the year, and the facets indicate the age group in years. Separate simulations are shown for each combination of parameters.

**Figure S4:**
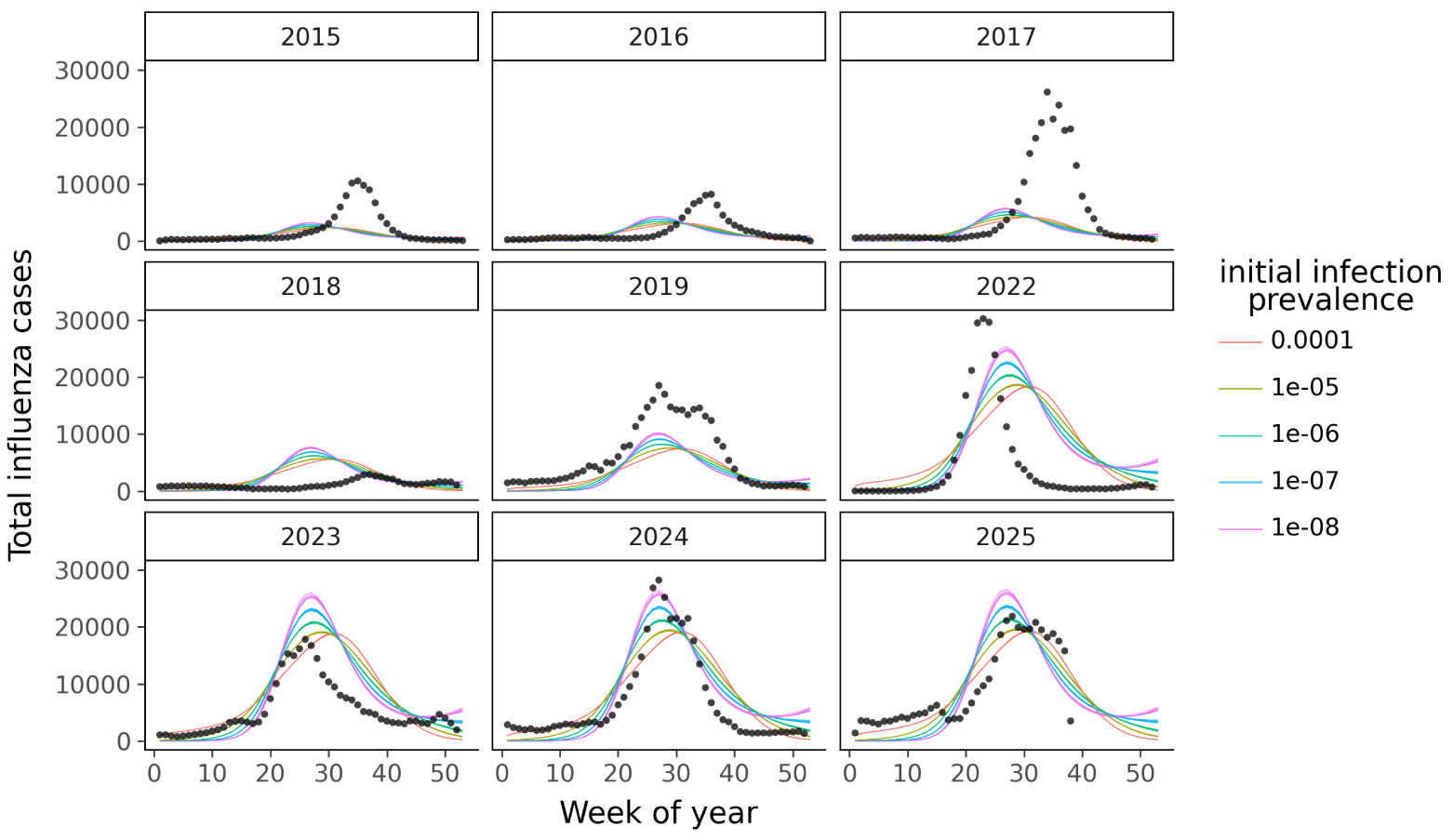
Model calibration of influenza cases by year for different initial infection prevalence. The number of cases are aggregated across all ages (black circles). The colour indicates the initial infection prevalence, and the facets indicate the year.

**Figure S5:**
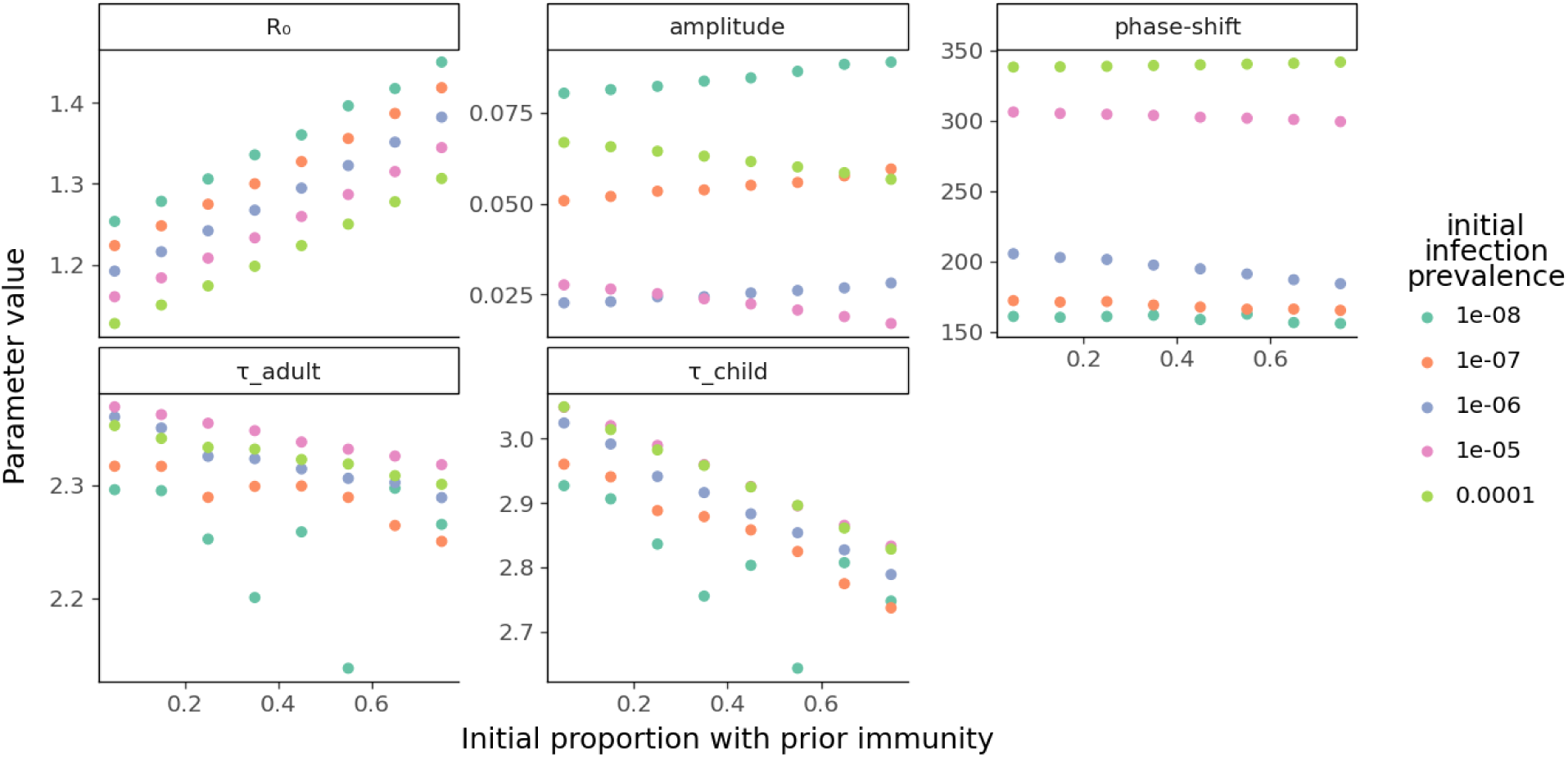
Maximum likelihood calibration values. The model was calibrated independently for various combinations of prior immunity and infection prevalence at the beginning of the simulations. The importance of seasonal forcing over a single year depended on the initial infection prevalence: the fitted amplitude varied between 0.02 and 0.09, with relatively low and high initial infection prevalence associated with a larger amplitude. For an initial infected proportion of 1E-7 and 1E-6 (orange and purple respectively), the fitted amplitude was very small, indicating the epidemic was largely driven by *R*_0_ and the initial proportion of partially susceptible individuals due to prior immunity.

**Figure S6:**
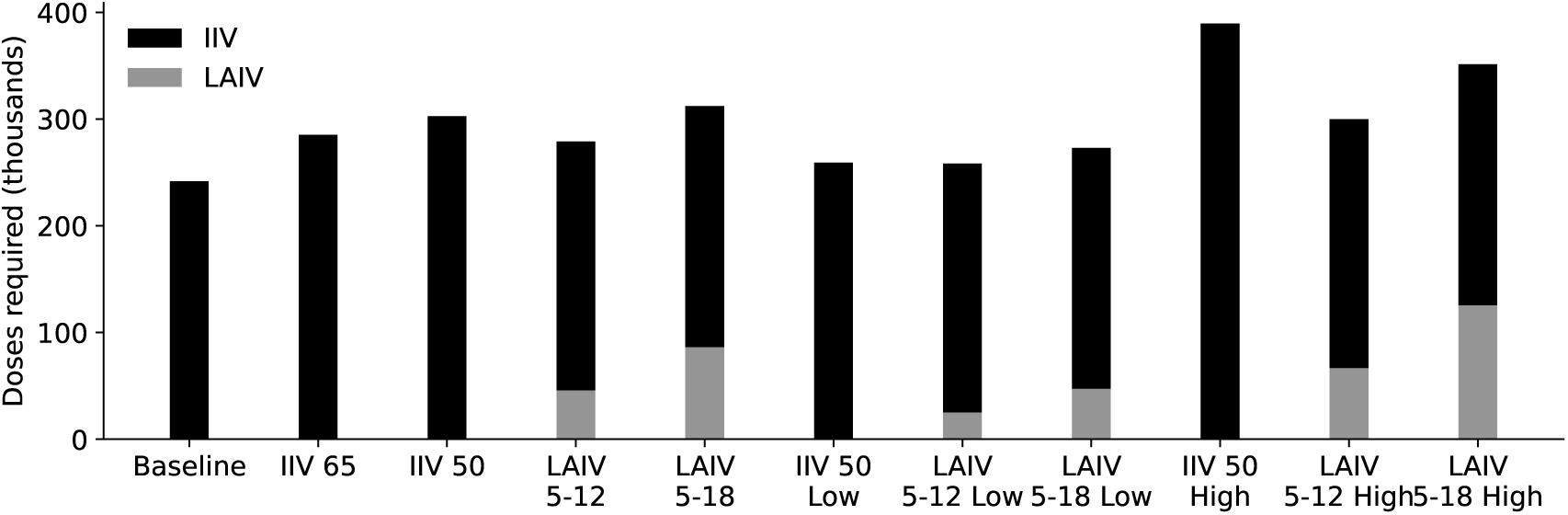
Modelled doses of inactivated influenza vaccine (IIV) and live-attenuated influenza vaccine (LAIV) used (per million population) by vaccine strategy (stacked).

**Figure S7:**
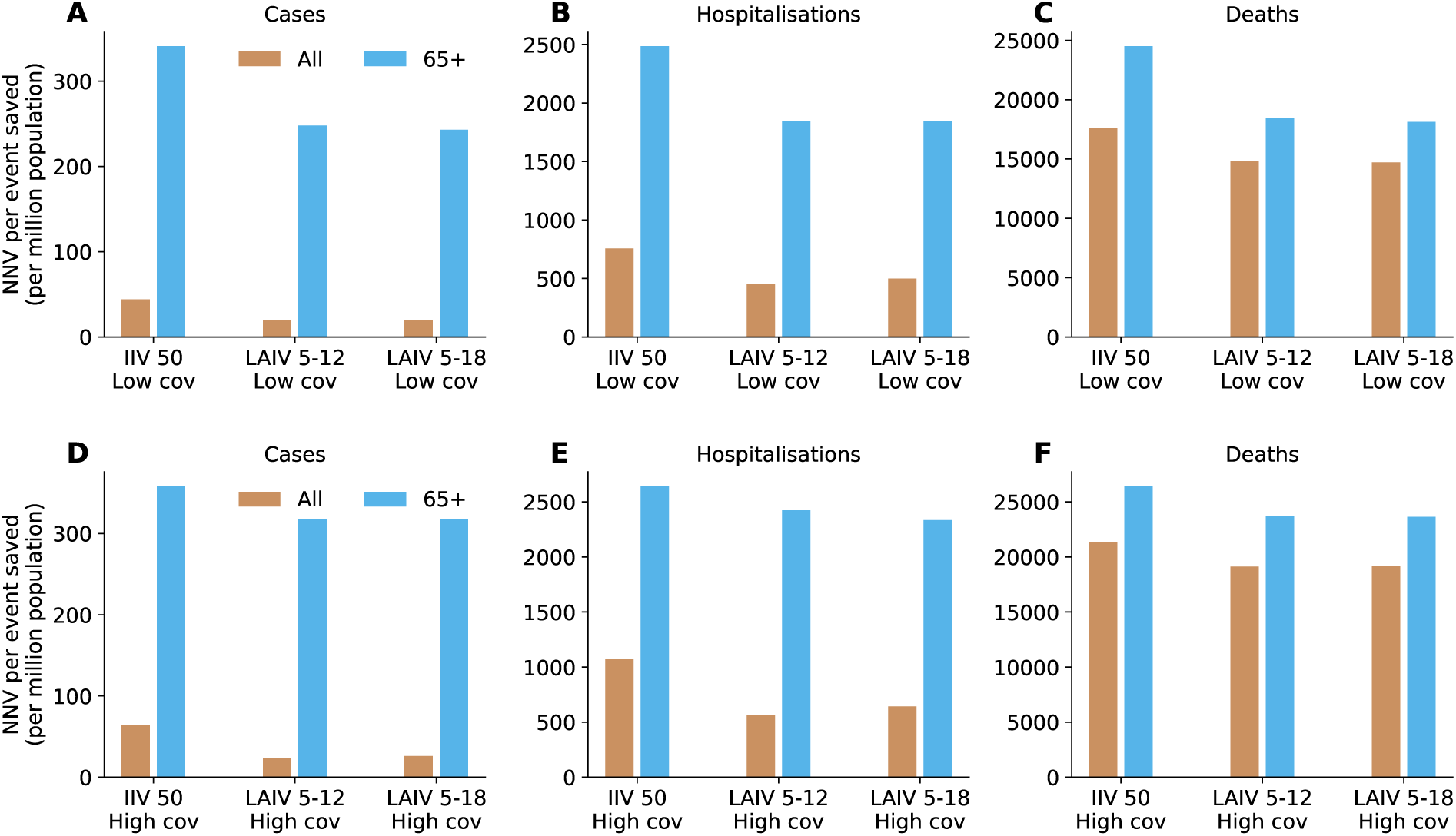
Mean number needed to vaccinate to prevent one case, hospitalisation, and death, compared to the baseline scenario, as part of sensitivity analysis. (A) Low coverage of 30% and (B) high coverage of 80% in targeted age groups. The bars represent the mean NNV over 40 simulations.

## B Supplementary Tables

**Table S1:**
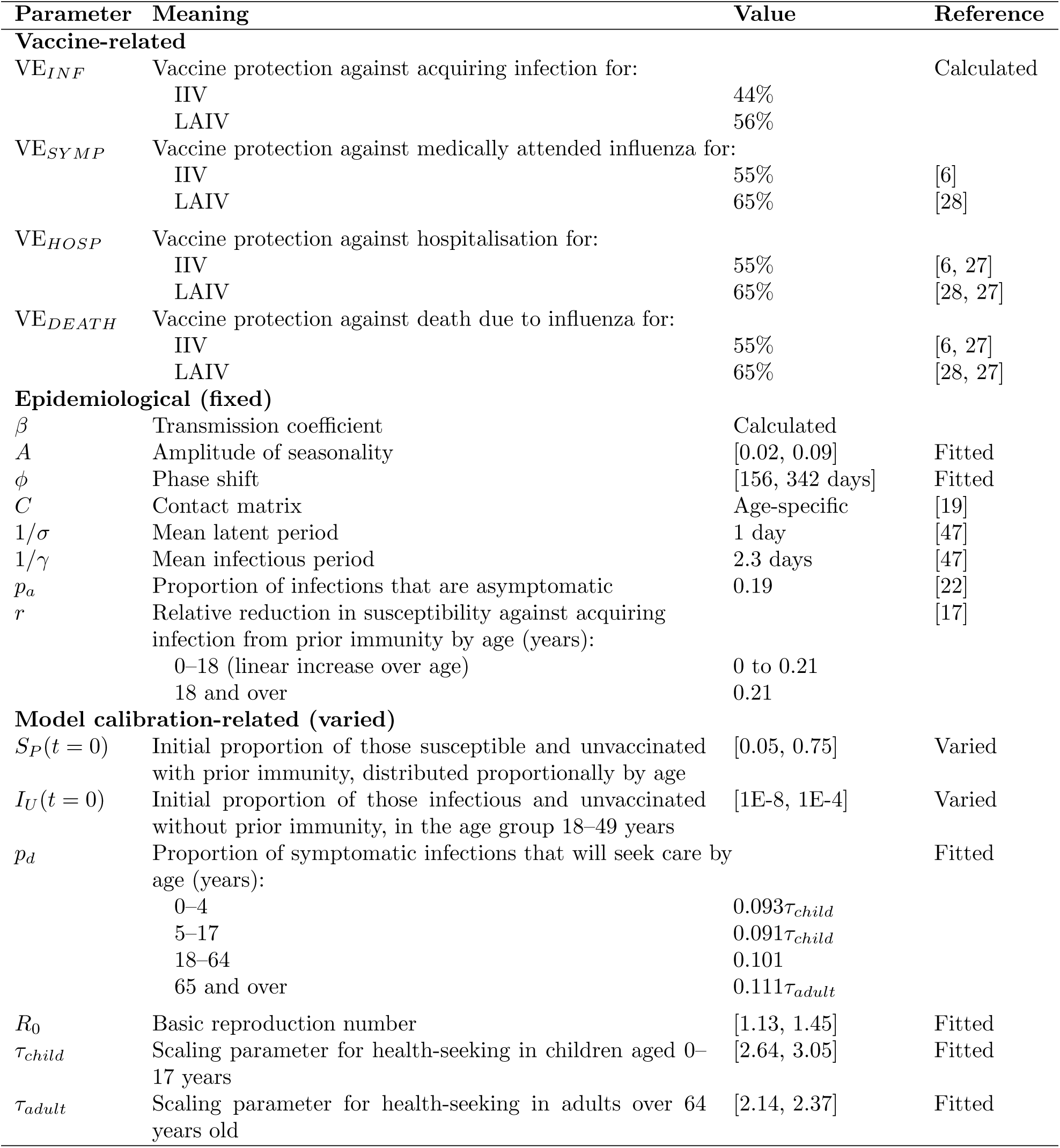
Descriptions and values for model parameters. Parameters are grouped into three broad categories: vaccine-related, epidemiological (fixed), and model calibration-related (varied). Where applicable, brackets show the range over 40 sets of initial population and fitted parameter values in the uncertainty analysis.

| Parameter | Meaning | Value | Reference |
|---|---|---|---|
| **Vaccine-related** |  |  |  |
| VE<sub>*INF*</sub> | Vaccine protection against acquiring infection for: |  | Calculated |
|  | IIV | 44% |  |
|  | LAIV | 56% |  |
| VE<sub>*SYMP*</sub> | Vaccine protection against medically attended influenza for: |  |  |
|  | IIV | 55% | [6] |
|  | LAIV | 65% | [28] |
| VE<sub>*HOSP*</sub> | Vaccine protection against hospitalisation for: |  |  |
|  | IIV | 55% | [6, 27] |
|  | LAIV | 65% | [28, 27] |
| VE<sub>*DEATH*</sub> | Vaccine protection against death due to influenza for: |  |  |
|  | IIV | 55% | [6, 27] |
|  | LAIV | 65% | [28, 27] |
| **Epidemiological (fixed)** |  |  |  |
| *β* | Transmission coefficient | Calculated |  |
| *A* | Amplitude of seasonality | [0.02, 0.09] | Fitted |
| *ϕ* | Phase shift | [156, 342 days] | Fitted |
| *C* | Contact matrix | Age-specific | [19] |
| 1/*σ* | Mean latent period | 1 day | [47] |
| 1/*γ* | Mean infectious period | 2.3 days | [47] |
| *p*<sub>*a*</sub> | Proportion of infections that are asymptomatic | 0.19 | [22] |
| *r* | Relative reduction in susceptibility against acquiring infection from prior immunity by age (years): |  | [17] |
|  | 0–18 (linear increase over age) | 0 to 0.21 |  |
|  | 18 and over | 0.21 |  |
| **Model calibration-related (varied)** |  |  |  |
| *S*<sub>*P*</sub>(*t* = 0) | Initial proportion of those susceptible and unvaccinated with prior immunity, distributed proportionally by age | [0.05, 0.75] | Varied |
| *I*<sub>*U*</sub>(*t* = 0) | Initial proportion of those infectious and unvaccinated without prior immunity, in the age group 18–49 years | [1E-8, 1E-4] | Varied |
| *p*<sub>*d*</sub> | Proportion of symptomatic infections that will seek care by age (years): |  | Fitted |
|  | 0–4 | 0.093*τ*<sub>*child*</sub> |  |
|  | 5–17 | 0.091*τ*<sub>*child*</sub> |  |
|  | 18–64 | 0.101 |  |
|  | 65 and over | 0.111*τ*<sub>*adult*</sub> |  |
| *R*<sub>0</sub> | Basic reproduction number | [1.13, 1.45] | Fitted |
| *τ*<sub>*child*</sub> | Scaling parameter for health-seeking in children aged 0–17 years | [2.64, 3.05] | Fitted |
| *τ*<sub>*adult*</sub> | Scaling parameter for health-seeking in adults over 64 years old | [2.14, 2.37] | Fitted |

**Table S2:**
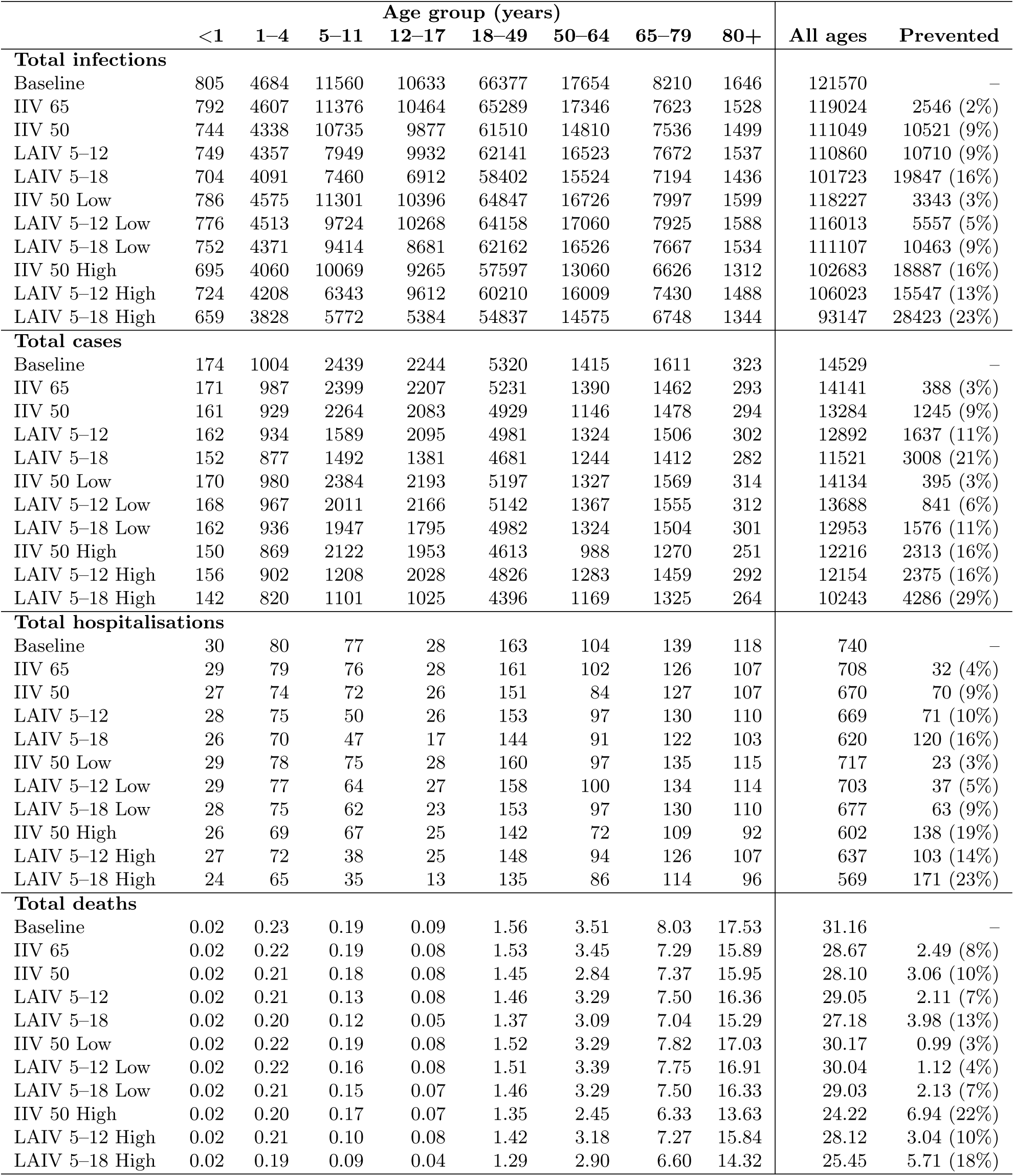
Total number of infections, cases, hospitalisations, and deaths per million population by vaccine strategy and scenario. The last column shows the absolute and relative reduction in prevented outcomes over all ages compared to baseline.

**Table S3:**
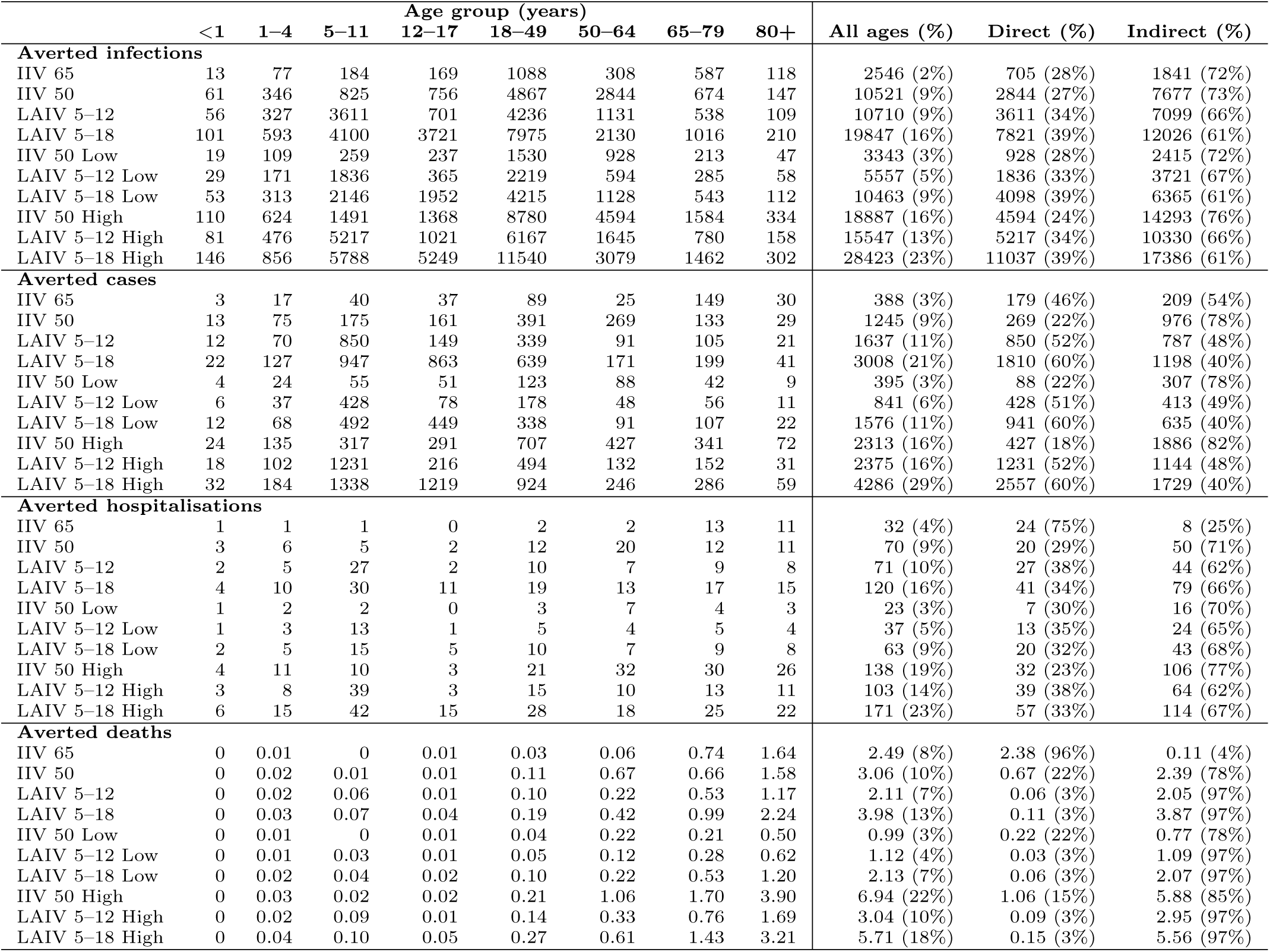
Averted infections, cases, hospitalisations, and deaths per million population by vaccine strategy (relative to baseline). The mean over 40 simulations is shown. The last two columns show the number of averted outcomes in the target age (direct) and outside of it (indirect), both in absolute numbers and percentages of the total outcomes averted.

**Table S4:**
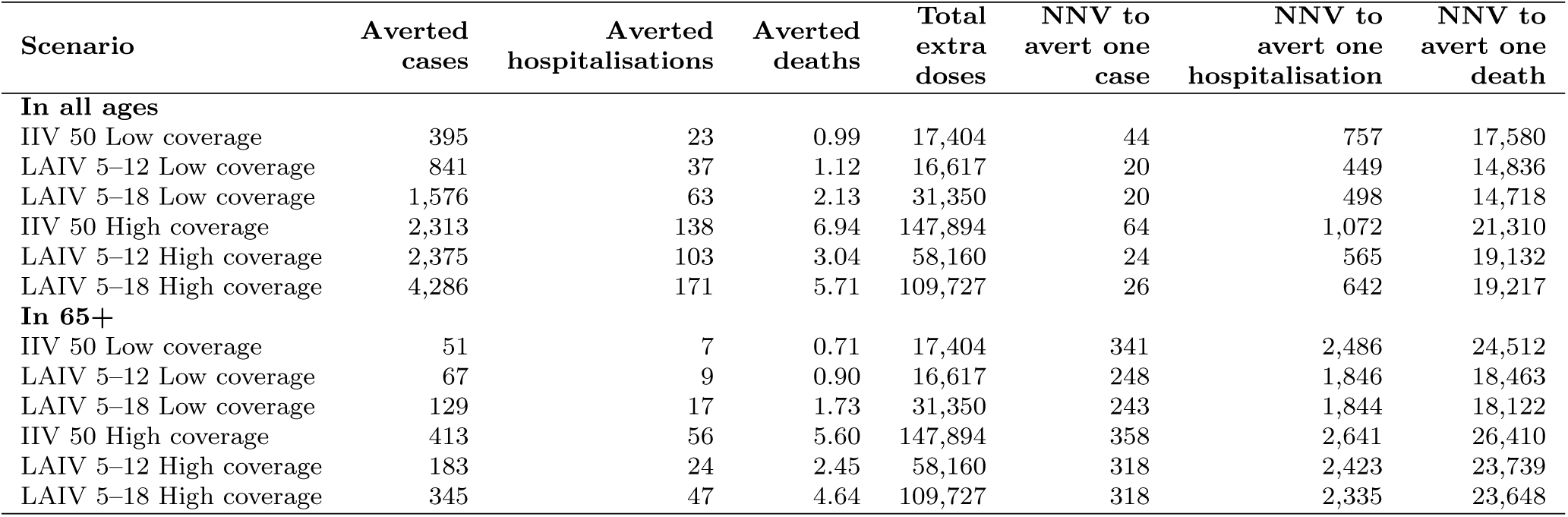
Number needed to vaccinate to avert 1 case, 1 hospitalisation, and 1 death in high coverage scenarios. (compared to baseline of 241,639 doses per million population), as part of a sensitivity analysis. The mean over 40 simulations is presented.

| **Scenario** | **Averted cases** | **Averted hospitalisations** | **Averted deaths** | **Total extra doses** | **NNV to avert one case** | **NNV to avert one hospitalisation** | **NNV to avert one death** |
|---|---|---|---|---|---|---|---|
| **In all ages** |  |  |  |  |  |  |  |
| IIV 50 Low coverage | 395 | 23 | 0.99 | 17,404 | 44 | 757 | 17,580 |
| LAIV 5–12 Low coverage | 841 | 37 | 1.12 | 16,617 | 20 | 449 | 14,836 |
| LAIV 5–18 Low coverage | 1,576 | 63 | 2.13 | 31,350 | 20 | 498 | 14,718 |
| IIV 50 High coverage | 2,313 | 138 | 6.94 | 147,894 | 64 | 1,072 | 21,310 |
| LAIV 5–12 High coverage | 2,375 | 103 | 3.04 | 58,160 | 24 | 565 | 19,132 |
| LAIV 5–18 High coverage | 4,286 | 171 | 5.71 | 109,727 | 26 | 642 | 19,217 |
| **In 65+** |  |  |  |  |  |  |  |
| IIV 50 Low coverage | 51 | 7 | 0.71 | 17,404 | 341 | 2,486 | 24,512 |
| LAIV 5–12 Low coverage | 67 | 9 | 0.90 | 16,617 | 248 | 1,846 | 18,463 |
| LAIV 5–18 Low coverage | 129 | 17 | 1.73 | 31,350 | 243 | 1,844 | 18,122 |
| IIV 50 High coverage | 413 | 56 | 5.60 | 147,894 | 358 | 2,641 | 26,410 |
| LAIV 5–12 High coverage | 183 | 24 | 2.45 | 58,160 | 318 | 2,423 | 23,739 |
| LAIV 5–18 High coverage | 345 | 47 | 4.64 | 109,727 | 318 | 2,335 | 23,648 |

## C Appendix

### Data

#### Influenza surveillance

Influenza is a notifiable disease in Australia. Notified influenza cases, disaggregated by age-group (*a*), notification week (*w*) and year (*y*), were obtained from the National Notifiable Disease Surveillance System (NNDSS) [21] from 2015 to 2025 (denoted *D_a,w,y_*) for all states in Australia. The NNDSS data only includes cases with laboratory confirmation. Age group was stratified by under 1, 1–4, 5–11, 12–17, 18–64, 65–79, and 80 years or older. We excluded data from years 2020 and 2021 due to the implementation of nonpharmaceutical interventions during the COVID pandemic and limited influenza transmission.

#### Test-seeking behaviour

To estimate the conditional probability that people take a test given symptomatic infection (*p_d_*) we used healthcare-seeking behaviour data from the FluTracking surveys; specifically, we calculated the proportion of reports of influenza symptoms where an influenza test was also sought, for each age group and for each year. We did not make any adjustment for the type of test (whether rapid antigen or polymerase chain reaction). FluTracking is a participatory influenza surveillance system in Australia and other participating regions, which includes regular surveys of volunteers with questions about symptoms and testing behaviour. FluTracking data for years 2015 to 2025 were analysed, and data from 2020 and 2021 were again excluded. The FluTracking surveys were available in age groups 0–4, 5–17, 18–64, and 65 years or older.

#### Vaccination

Influenza vaccination coverage data were obtained from the National Centre for Immunisation Research and Surveillance (NCIRS) from 2021 to 2024 [7]. Data on weekly cumulative vaccination coverage were available from the beginning of March to the end of August in each year, spanning notification weeks 10 to 34. Data were disaggregated into 5 age groups (6 months to 4 years; 5 to 14 years; 15 to 49 years; 50 to 64 years; and 65 years or older).

#### Influenza hospitalisations

We obtained principal diagnosis data for the years 2018–19, 2022–23, and 2023–24 from the Australian Institute of Health and Welfare National Hospital Morbidity Database [25]. The data contains separation counts from public and private Australian hospitals for all states and territories. Counts were disaggregated into 19 age groups (under 1 year, 1–4 years, 5–84 years in 5-year bins, and 85+ years) from July to June for multiple years. We used population sizes for each corresponding year from the Australian Bureau of Statistics (ABS) [18] and calculated the hospitalisations per population for our modelled age groups. We calculated the age-specific and all-population average influenza hospitalisation rates across the three one-year periods and used this average in our analysis (Figure S2A).

#### Influenza deaths

We obtained data on age-specific deaths due to influenza in Australia, coded to J09–J11, for the years 2023, 2024, and 2025 from the ABS [4]. Because in the ABS data, influenza deaths in children are reported as one age group (0–19 years), we further stratified the deaths in this group to align with our modelled age groups, using the age-specific proportions of influenza deaths in child age groups across the years 2018–2023 reported in McRae et al. 2025 [48]. We then calculated the age-specific and all-population average influenza death rates across the three years and used this average in our analysis (Figure S2B).

### Model description

We developed an age-structured, deterministic compartmental model of seasonal influenza transmission and vaccination. In the model, the population is divided into 9 age groups: 0–*<*0.5, 0.5–*<*1, 1–4, 5–11, 12–17, 18–49, 50–64, 65–79, and 80+ years. The population is divided by infection status (susceptible, exposed, infectious, and recovered, represented by state variables *S, E, I, R* respectively) and vaccination status (unvaccinated without prior immunity, unvaccinated with prior immunity, and vaccinated, denoted by subscripts *U, P, V* respectively). Susceptible individuals become exposed but not yet infectious. After infection, they recover and become fully immune. Those who are vaccinated or have prior immunity gain a reduced susceptibility to acquiring infection by a proportional factor (1 *−* VE*_INF_*) and (1 *− r*) respectively. We use a gamma-distributed latent period, leading to a two-step transition from *E* to *I* [49]. Those who are vaccinated or unvaccinated with prior immunity have a reduced susceptibility to infection by a proportional factor (1 *− V E_INF_*) and (1 *− r*) respectively.

The model equations are:

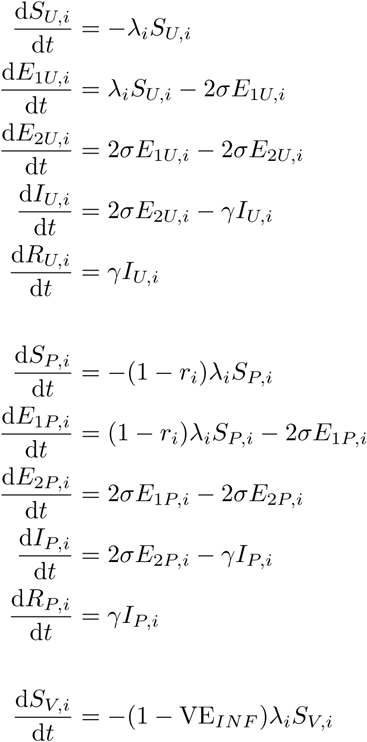

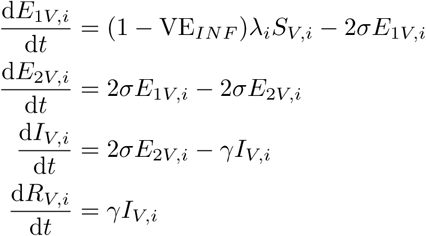

for age group *i* = 1, 2, 3*, …,* 9. The force of infection is

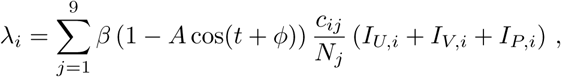

where *β* is the transmission coefficient; *c_ij_* is the average number of contacts per day between age classes *i* and *j*; *A* is the amplitude of seasonal forcing; and *ϕ* is the phase shift. The transmission coefficient was calculated with the next-generation matrix using a fitted *R*_0_. The model diagram is in the main text. Table S1 shows the model parameters and values.

#### Vaccine rollout

The rollout of both vaccines began in Week 10, with the IIV rollout taking place over 24 weeks and the LAIV rollout, over 12 weeks. We assumed a constant two-week delay from vaccine delivery to protection for both vaccine types. We vaccinated only susceptible or recovered individuals. We used a ‘leaky’ vaccine that provided partial protection against infection and hospitalisation to all vaccinated individuals [29]. We assumed that the vaccine had no effect on infectiousness in those who experienced breakthrough infection.

The weekly rollout rate of each vaccine was determined by the overall vaccination coverage of the age group (Table 1) and the overall speed of delivery of that vaccine. The IIV delivery speed varied weekly and was determined using cumulative influenza vaccination coverage data in two steps. First, we aggregated the data by age group for each year. Second, we normalised the weekly rate by the cumulative total of that year and set the mean per week as the overall speed of IIV delivery. The speed of LAIV delivery was assumed to be uniform over the LAIV rollout.

### Model calibration

We used FluTracking data [23] to inform the assumed age-specific fractions of infections that seek medical care. We modelled the proportion of people with symptoms who sought care prior to 2020 using a logistic regression model (*p_ft_*) and the mean values for all years post 2021. The modelled increase in healthcareseeking behaviour from 2022 onwards captures differences in test-seeking and structural changes in question design following the pandemic. The age group (categorical) and year (continuous) were included as covariates in the logistic regression. A comparison of people seeking care for COVID-19 in the FluTracking data and a weekly, national behavioural survey established by the Australian government found test-seeking behaviour among those with symptoms was higher in the FluTracking participants (84.9%) compared to those in the national behavioural survey (58.8%) [24], therefore we scaled the estimated proportion of people seeking care in the FluTracking data by 58.8*/*84.9 (Figure S1). *p_d_* in children and adults *≥*65 years old was informed during the model calibration by scaling the *p_ft,a_* values thus,

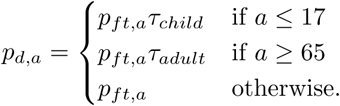

The model was calibrated to case notification data aggregated across all Australian states and territories. For the calibration we used the year-specific Australian population size [18]. The model was calibrated with a fixed proportion of the population with prior immunity and initial seed prevalence on the first week. We conducted a sensitivity analysis of these parameters, such that we started the model simulations with different proportions of susceptible and unvaccinated population. Results are presented over a one-year time horizon from January to December, where a single LAIV campaign is implemented before the winter season.

In all simulations we ran the model over a single seasonal epidemic and calculated the weekly number of cases with the status quo vaccination schedule. We calibrated the model by allowing the (1) *R*_0_, (2) amplitude of seasonal forcing (*A*), (3) phase shift of seasonal forcing (*ϕ*), and (4) scaling factors (*τ_child_* and *τ_adult_*) to adjust the proportion of infections that seek a test (*p_ft_*) to vary. Infections were seeded at the beginning of the year. We assumed that cases are Poisson distributed, which assumes the variance is equal to the mean, and calculated the probability mass function thus,

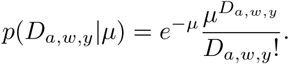

The log-likelihood of observing the NNDSS data was calculated thus,

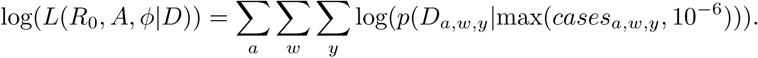

Note that we specified a weekly minimum number of infections of 10*^−^*^6^.

The sensitivity of the model calibration to different numbers of people with prior immunity and infection prevalence at the beginning of the year was investigated by calibrating the model with the initial proportion of those with prior immunity (*S_P_*) ranging from 5% to 75% distributed proportionally by age, which was determined by the approximate range of the proportion of people with Hemagglutination-Inhibition titre greater than 40 observed for H3N2 influenza in Australia [50]. People in the *S_P_* compartment had reduced susceptibility to infection (given by the *r* parameter; Table S1) relative to those without prior immunity. The initial infection prevalence ranged from 1E-8 to 1E-4.

### Model outcomes of interest

To calculate the age-specific incidence, *inc*, we tracked the vaccination and age-specific total number of recovered infections (*T*):

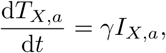

where *X ∈ U, V, P*. We calculated the case incidence at each time step as,

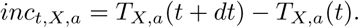

The weekly incidence was calculated as,

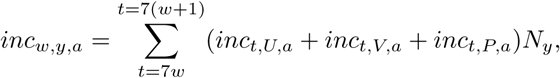

where *N_y_*is the population size of Australia for the given year for the model calibration or 1 million for the scenario modelling.

We used three main model outcomes of interest to evaluate the performance of each vaccine strategy.

They are

- weekly incident infections
- weekly incident cases of disease
- weekly incident hospitalisations due to influenza, calculated by

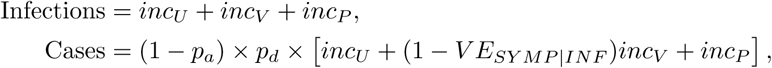

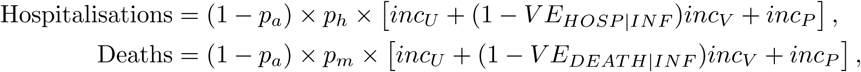

where *inc_X_* is the incidence for those who are unvaccinated without prior immunity, vaccinated, and unvaccinated with prior immunity (*X* = *U, V, P*, respectively); (1 *− p_a_*) is the proportion of infections that is symptomatic; *p_d_* is the proportion of symptomatic infections that will seek medical care; *p_h_* is the proportion of symptomatic infections that will require hospitalisation; and *p_m_* is the proportion of symptomatic infections that will result in death. The age-specific proportions *p_h_* and *p_m_* were calculated from data on influenza hospitalisations [25] and on influenza mortalities [4, 46], respectively, and modelled baseline infections. Vaccinated individuals gain partial protection to medically attended influenza (*V E_SY MP_*),influenza hospitalisation (*V E_HOSP_*), and death due to influenza (*V E_DEAT H_*), from which the vaccine effects *conditional on becoming infected* [26] were calculated thus:

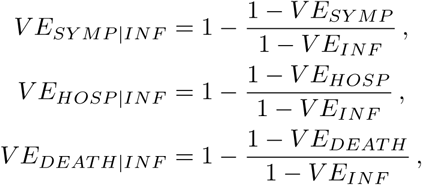

where *V E_INF_*is the vaccine effectiveness against acquiring infection.

